# Divergent SARS CoV-2 Omicron-specific T- and B-cell responses in COVID-19 vaccine recipients

**DOI:** 10.1101/2021.12.27.21268416

**Authors:** Corine H. GeurtsvanKessel, Daryl Geers, Katharina S. Schmitz, Anna Z. Mykytyn, Mart M Lamers, Susanne Bogers, Lennert Gommers, Roos S.G. Sablerolles, Nella N. Nieuwkoop, Laurine C. Rijsbergen, Laura L.A. van Dijk, Janet de Wilde, Kimberley Alblas, Tim I. Breugem, Bart J.A. Rijnders, Herbert de Jager, Daniela Weiskopf, P. Hugo M. van der Kuy, Alessandro Sette, Marion P.G. Koopmans, Alba Grifoni, Bart L. Haagmans, Rory D. de Vries

## Abstract

The severe acute respiratory distress syndrome coronavirus-2 (SARS-CoV-2) Omicron variant (B.1.1.529) is spreading rapidly, even in vaccinated individuals, raising concerns about immune escape. Here, we studied neutralizing antibodies and T-cell responses to SARS-CoV-2 D614G (wildtype, WT), and the B.1.351 (Beta), B.1.617.2 (Delta), and B.1.1.529 (Omicron) variants of concern (VOC) in a cohort of 60 health care workers (HCW) after immunization with ChAdOx-1 S, Ad26.COV2.S, mRNA-1273 or BNT162b2. High binding antibody levels against WT SARS-CoV-2 spike (S) were detected 28 days after vaccination with both mRNA vaccines (mRNA-1273 or BNT162b2), which significantly decreased after 6 months. In contrast, antibody levels were lower after Ad26.COV2.S vaccination but did not wane. Neutralization assays with authentic virus showed consistent cross-neutralization of the Beta and Delta variants in study participants, but Omicron-specific responses were significantly lower or absent (up to a 34-fold decrease compared to D614G). Notably, BNT162b2 booster vaccination after either two mRNA-1273 immunizations or Ad26.COV.2 priming partially restored neutralization of the Omicron variant, but responses were still up to-17-fold decreased compared to D614G. CD4+ T-cell responses were detected up to 6 months after all vaccination regimens; S-specific T-cell responses were highest after mRNA-1273 vaccination. No significant differences were detected between D614G- and variant-specific T-cell responses, including Omicron, indicating minimal escape at the T-cell level. This study shows that vaccinated individuals retain T-cell immunity to the SARS-CoV-2 Omicron variant, potentially balancing the lack of neutralizing antibodies in preventing or limiting severe COVID-19. Booster vaccinations may be needed to further restore Omicron cross-neutralization by antibodies.

## Introduction

The severe acute respiratory distress syndrome coronavirus-2 (SARS-CoV-2) Omicron variant (B.1.1.529) is characterized by a high number of mutations in the spike (S) protein that have immune evasive potential. Based on transmission characteristics and immune evasion, the World Health Organization (WHO) designated Omicron as a novel variant of concern (VOC). The Omicron variant has been identified worldwide and models have predicted rapid surges of cases that could surpass earlier peaks (*1*). More data are needed to understand the Omicron disease severity profile, and how severity is impacted by vaccination and pre-existing immunity (*2*).

The large number of mutations and deletions in the Omicron S protein include alterations in the receptor binding domain (RBD), the main target of neutralizing antibodies responsible for host cell entry (*3-5*). It was previously shown for the Beta (B.1.351) and Omicron variant that mutations within the RBD (*6-8*) and the N-terminal domain (*9*) can lead to partial escape from neutralizing antibodies. Preliminary data indeed show that there is a concerning reduction in neutralizing antibody titers against Omicron compared to D614G in convalescent and vaccinated individuals, which could be partially restored by booster vaccination (*10-12*). Additionally, initial show that Omicron is resistant to most antibodies authorized for clinical use (*13*).

Thus far, neutralizing antibodies are regarded the main correlate of protection against SARS-CoV-2 infection (*14, 15*). The relative contribution of virus-specific T-cells is more difficult to decipher. SARS-CoV-2-specific T-cells clear infected cells, thereby contributing to the reduction of viral replication, potentially limiting pathogenicity (*16*). Previous studies on the impact of mutations in the S protein on T-cell recognition show that VOC S proteins are equally recognized by S-specific T-cells induced by mRNA-(*8, 17, 18*) and adenovirus-based (*19*) vaccines, but studies on T-cell recognition of Omicron are scarce up to present(*20*). The numerous mutations in the S protein in the Omicron variant potentially disrupt T-cell epitopes (*21*); however, this negative impact is likely to be significantly smaller than the effect on neutralizing antibody epitopes.

Five vaccines have now been authorized for use in Europe by the European Medicines Agency (EMA); mRNA-based (mRNA-1273 [Moderna], BNT162b2 [Pfizer]), adenovirus vector-based (ChAdOx-1 S [Astrazeneca], Ad26.COV2.S [Janssen]) and the protein-based NVX-CoV2373 (Novavax). These vaccines all use the SARS-CoV-2 S protein of the ancestral strain as template for design, induce robust immune responses, and protect from developing severe coronavirus disease-2019 (COVID-19) (*22-25*). However, vaccine efficacy differs and may be affected by waning of antibodies and the emergence of variants (*15, 26-28*). In general, boosting immune responses by additional vaccinations efficiently restores antibody and T-cell responses, but clinical efficacy and protection against Omicron remains to be determined.

We analysed humoral and cellular immune responses early and late (up to 6 months) after vaccination with ChAdOx-1, S, Ad26.COV2.S, mRNA-1273 or BNT162b2 and performed in-depth analyses of cross-reactivity of neutralizing antibodies and T-cells against against the D614G (wildtype, WT), Beta, Delta and Omicron variants. Additionally, we assessed cross-recognition of variants by neutralizing antibodies and T-cells after booster vaccination. We found cross-neutralization of the Delta and Beta variant in all vaccinated and convalescent individuals, but neutralization of Omicron was consistently lower or even absent. Strikingly, all VOC were equally recognized by CD4+ T-cells. Booster vaccination restored Omicron-specific neutralization, but still at significantly lower levels compared to D614G.

## Results

### Cohort description

To assess binding antibody and T-cell responses after different vaccination regimens, we collected serum and PBMC from N=400 participants, of which N=26 received two doses ChAdOx-1 S, N=75 received a single dose Ad26.COV2.S, N=199 received 2 doses of mRNA-1273, and N=100 received 2 doses of BNT162b2. Additionally, we measured responses in N=23 plasma donors with a confirmed WT SARS-CoV-2 infection. Binding antibodies were assessed early (28 days after BNT162b2, mRNA-1273 or ChAdOx-1, or 56 days after Ad26.COV2.S) and late (6 months) after vaccination. T-cell responses in whole-blood were assessed at 56 days and 6 months after vaccination (Ad26.COV2.S, N=31), 28 days and 6 months after second vaccination (mRNA-1273, N=39), or exclusively at 6 months after vaccination (ChAdOx-1 S, N=14). Virus neutralization and T-cell assays against VOC were performed in a selection of N=15 participants from each vaccination group at the early and late timepoint. The study design is shown in **Figure 1A**; participant characteristics are summarized in **Table 1**.

**Figure 1.**
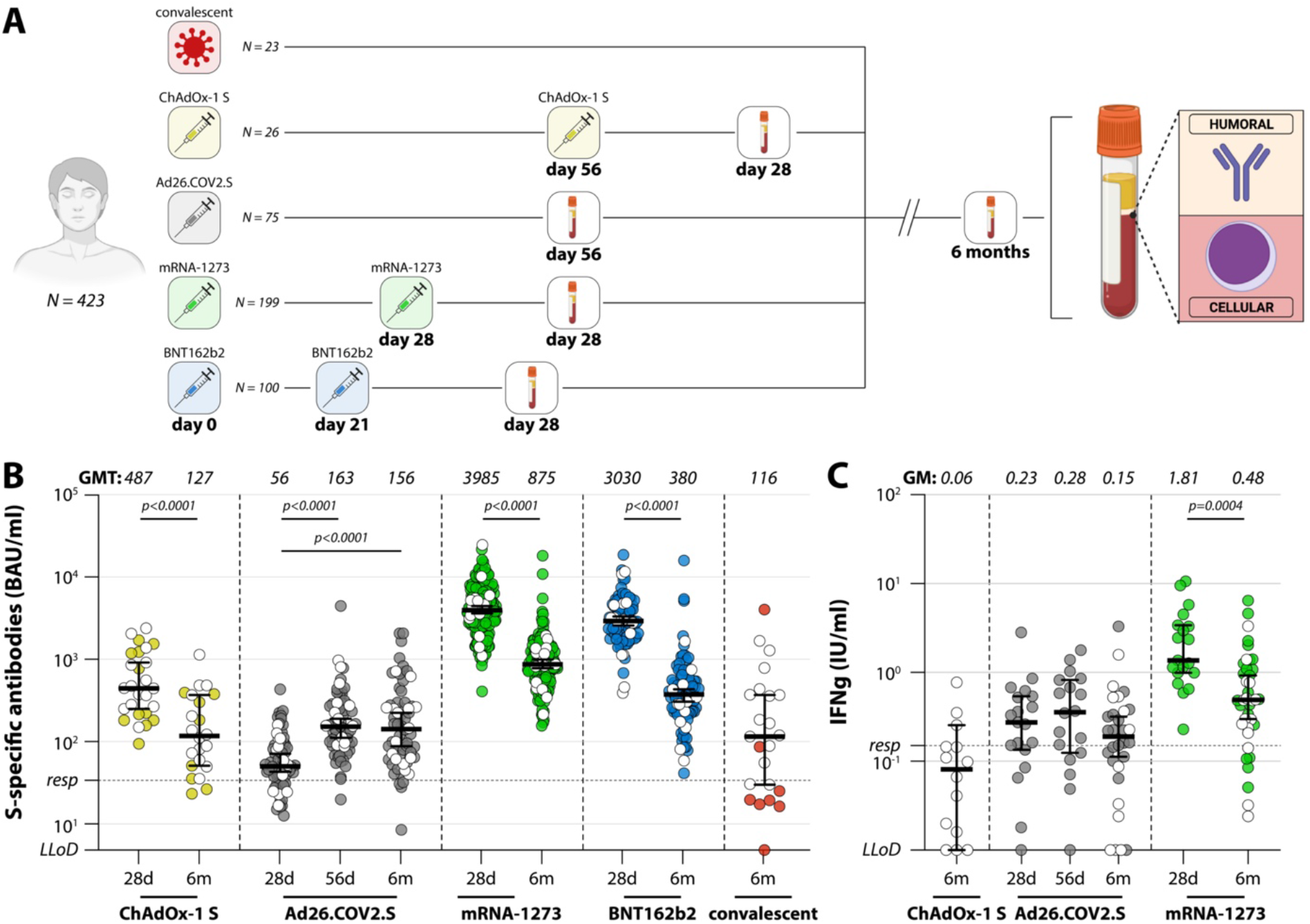
Study design and detection of virus-specific binding antibodies and T-cells. (A) Diagram of the number of included participants and study groups. A total of N=423 participants were included for the analysis of binding antibodies and T-cell responses, responses were measured early and late after completion of the vaccination regimen. Binding antibodies were also assessed in N=23 convalescent participants. (B) Levels of binding S-specific antibodies early (28 days after second vaccination, or 56 days after Ad26.COV2.S vaccination) and late (6 months after completion of vaccination regimen) after vaccination or infection. LLoD is 4.81 BAU/ml, responder (resp) cut-off is 33.8 BAU/ml (dotted line). Geometric mean titers are indicated above the graph. (C) IFNγ levels in plasma after stimulation of whole-blood with peptide pools spanning the S protein (Ag2, QIAGEN) early and late after ChAdOX-1 S, Ad26.COV2.S, or mRNA-1273 vaccination, ChAdOx-1 S responses were exclusively measured at 6 months). LLoD is 0.01 IU/ml, responder cut-off is 0.15 IU/ml. Geometric means are indicated above the graph. Comparisons of timepoints within study groups were performed by paired T test. White symbols represent samples selected for in-depth analyses. LLoD = lower limit of detection, S = Spike, BAU = binding arbitrary units, GMT = geometric mean titer, d = days, m = months, IFNγ = interferon gamma.

**Table 1.** Participant characteristics.

| Cohort | N | Age (median years, 95% CI) | Female (N, %) | Vaccination interval<br>(median days, 95% CI) |
| --- | --- | --- | --- | --- |
| Convalescent | 23 | 48 (42-58) | 4 (17%) | N/A |
| *In-depth* | 15 | 53 (42-62) | 1 (7%) | N/A |
| ChAdOx-1 S | 26 | 63 (62-64) | 17 (61%) | 56 (56-70) |
| *In-depth* | 15 | 63 (62-64) | 9 (60%) | 56 (56-70) |
| Ad26.COV2.S | 75 | 37 (32-43) | 62 (83%) | N/A |
| *In-depth* | 15 | 38 (32-50) | 13 (87%) | N/A |
| mRNA-1273 | 199 | 41 (39-44) | 156 (78%) | 28 (28-28) |
| *In-depth* | 15 | 34 (27-40) | 12 (80%) | 25 (25-28) |
| BNT162b2 | 100 | 40.5 (37-45) | 66 (66%) | 21 (21-21) |
| *In-depth* | 15 | 46 (36-53) | 11 (73%) | 22 (21-25) |

Participants in the ChAdOx-1 S group were significantly older than participants in other groups (p<0.0001, Kruskal-Wallis with multiple comparisons); other groups were comparable. Additionally, the intervals between first and second vaccination were different for all groups that received two vaccines: median interval for (1) ChAdOx-1 S was 56 days, (2) mRNA-1273 was 28 days, and (3) BNT162b2 was 21 days (p<0.0001 for all comparisons, Kruskal-Wallis with multiple comparisons).

### Vaccine-induced antibodies wane after 6 months, except in Ad26.COV2.S vaccinated HCW

Highest levels of S-specific binding antibodies were detected 28 days after full vaccination with mRNA-1273 or BNT162b2 (geometric mean titer [GMT] of 3985 and 3030 BAU/ml, respectively). A significant reduction in GMT was observed after 6 months in both groups (**Figure 1B**, p<0.0001, paired T test). Both adenovirus vector-based vaccines induced significantly lower binding antibody titers at 28 days (ChAdOx-1 S) or 56 days (Ad26.COV2.S) after vaccination (GMT of 127 and 163 BAU/ml, p<0.0001, unpaired T test for all comparisons). Whereas a significant drop in antibody levels was observed in ChAdOx-1 S-vaccinated HCW (p<0.0001, paired T test), the levels remained stable in Ad26.COV2.S-vaccinated HCW. Despite the levels being stable in Ad26.COV2.S-vaccinated HCW, after 6 months the antibody levels were still significantly higher in HCW vaccinated with an mRNA-based vaccine (p<0.0001, unpaired T test Ad26.COV2.S versus both mRNA-1273 and BNT162b2). Interferon (IFN)γ levels were analyzed in ChAdOx-1 S-, Ad26.COV2.S, and mRNA-1273-vaccinated participants after stimulation of whole-blood as a measure for T-cell activity. mRNA-1273-vaccinated HCW had the highest T-cell responses, but significant decreases were observed 6 months after vaccination (p=0.0004, paired T test). T-cell responses remained stable up to 6 months after Ad26.COV2.S vaccination (**Figure 1C**).

### Significant reduction in Omicron neutralization in convalescent and vaccinated participants

Antibody functionality was measured in an infectious virus neutralization assay with passage 3 SARS-CoV-2 viruses D614G (WT), B.1.351 (Beta), B.1617.2 (Delta) and B.1.1.529 (Omicron) (**Figure 2A**). Human airway Calu-3 cells were used for virus propagation and neutralization assays because SARS-CoV-2 enters these cells using the TMPRSS2-mediated entry pathway (*29-32*). This entry pathway is used in vivo, and prevents adaptations in S, commonly observed in Vero cells. At 28 days post vaccination, mRNA-1273 vaccination elicited highest PRNT50 against D614G and all variants, followed by BNT162b2, ChAdOx-1 S, and Ad26.COV2.S vaccination. Delta and Beta variants were neutralized at slightly lower PRNT50 (especially after vaccination with mRNA-based vaccines) and a significant reduction of PRNT50 to Omicron was observed in all groups (p<0.0001 for ChAdOx-1 S, BNT162b2, and mRNA-1273, p=0.0001 for Ad26.COV2.S, Friedman test with multiple comparisons). Notably, 13/15 participants in the Ad26.COV2.S group did not neutralize the Omicron variant (**Figure 2C**).

**Figure 2.**
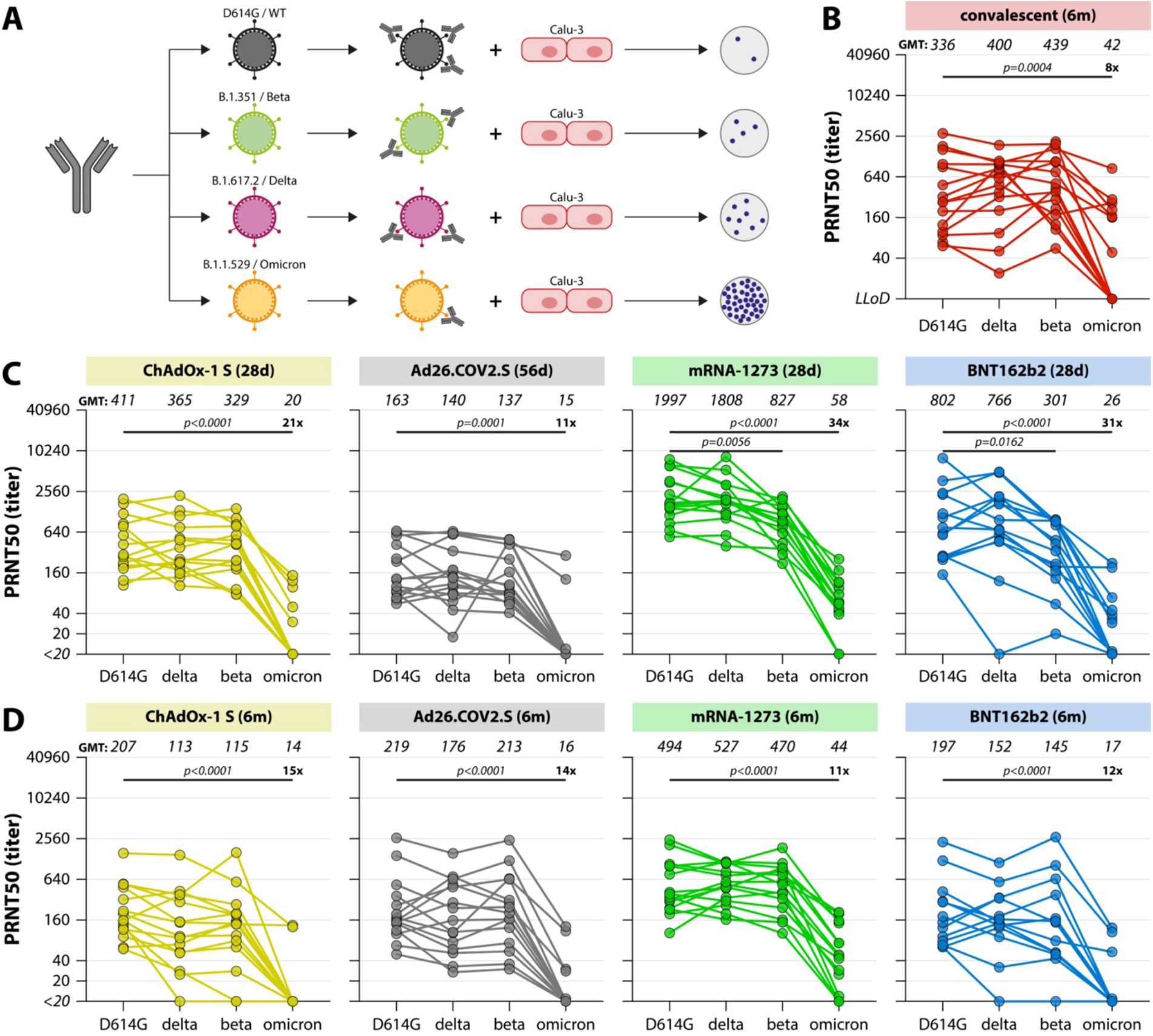
SARS-CoV-2 variant-specific neutralizing antibody responses. (A) Schematic overview of the infectious virus PRNT50 assay. Serum samples were pre-incubated with 400 plaque forming units infectious virus for 1 hour, transferred to Calu-3 cells, and N-positive plaques were counted after 8 hours. A PRNT50 (50% reduction of plaques) was calculated by a proportionate distance formula. (B, C, D) Levels of neutralizing antibodies in convalescent donors after 6 months (B), early after completion of the vaccination regimen (C), and late after completion of the vaccination regimen (D). The lowest serum dilution tested was 1:20, undetectable PRNT50 values (<20) were set at a PRNT50 of 10. Geometric mean titers are indicated above the graphs, fold change reductions are indicated and calculated by dividing the D614G GMT by the Omicron GMT. Comparisons of VOC-specific responses within study groups were performed by Friedman test with multiple comparisons. WT = wildtype, d = days, m = months, PRNT50 = plaque reduction neutralization titer – 50%,

At 6 months post vaccination, neutralizing antibodies against D614G waned after ChAdOx-1 S, mRNA-1273 and BNT162b2 vaccination, but remained stable after Ad26.COV2.S vaccination (**Figure 2C, 2D**). Differences in GMT PRNT50 values for the different vaccine groups were smaller at this point (GMT 207 for ChAdOx-1, 219 for Ad26.COV2.S, 494 for mRNA-1273, and 197 for BNT162b2). An 11 to 15-fold reduction in PRNT50 to Omicron led to low or completely absent neutralizing antibody levels in all groups (p<0.0001, Friedman test with multiple comparisons). An 8-fold reduction in Omicron neutralization was observed in the selection of 15 convalescent (after WT SARS-CoV-2 infection) sera p=0.0004, Friedman test with multiple comparisons, **Figure 2B**). Neutralizing antibodies to the D614G, Beta and Delta variants correlated well to measured binding antibody levels, whereas a lower (but significant) correlation was observed between Omicron-specific neutralizing antibodies and binding antibody levels (**Supplemental Figure 1**).

### Vaccination-induced S-specific CD4+ T-cells equally recognized VOC including Omicron

Besides neutralizing antibodies, we assessed the presence of S-specific T-cell responses by activation-induced marker (AIM) assay in the selection of participants for in-depth assays early and late after vaccination (**Figure 3**). For the selection of BNT162b2-vaccinated individuals a limited set of PBMC was available (N=5 out of 15). Peripheral blood mononuclear cells (PBMC) were stimulated with either overlapping peptide pools representing the full-length WT S protein, or peptide pools based on the S proteins of the Beta, Delta or Omicron variants (**Figure 3A**). After stimulation, AIM (OX40 and CD137) expression within CD4+ T-cells was measured by flow cytometry (gating strategy shown in **Figure 3B**).

**Figure 3.**
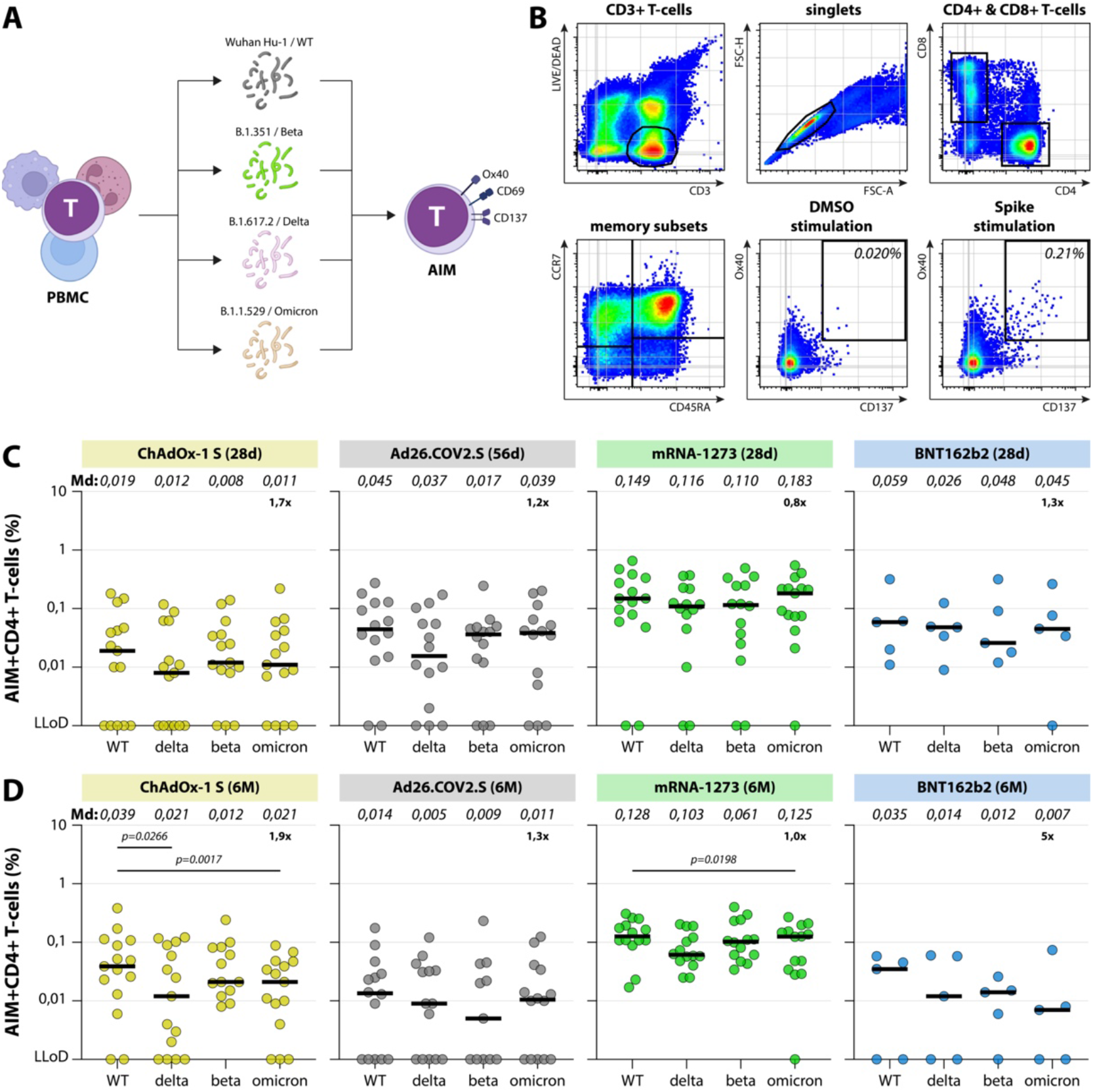
SARS-CoV-2 variant-specific T-cell responses. (A) Schematic overview of AIM assay. PBMC were stimulated with different overlapping peptide pools for 20 hours, followed by measurement of upregulation of activation markers by flow cytometry. (B) SARS-CoV-2-specific T-cells were detected by flow cytometry. After setting a time gate, LIVE CD3+ T-cells were gated, singlets were selected, and T-cells were subtyped into CD3+CD4+ and CD3+CD8+ cells. Within the CD4+ and CD8+ T-cells, T_naive_ were defined as CD45RA+CCR7+, T_CM_ as CD45RA-CCR7+, T_EM_ as CD45RA-CCR7-, and T_EMRA_ as CD45RA+CCR7-. S-specific T-cells were detected by co-expression of OX40 and CD137 on CD4+ T-cells in the combination of memory subsets. (C, D) Upregulation of AIM early after completion of the vaccination regimen (C) and late after completion of the vaccination regimen (D). Percentages indicate the percentage of CD4+OX40+CD137+ cells after subtraction of observed background in a DMSO stimulation. Geometric means are indicated above the graphs, fold change reductions are indicated and calculated by dividing the WT Md by the Omicron Md. Comparisons of VOC-specific responses within study groups were performed by Friedman test with multiple comparisons. Md = median, PBMC = peripheral blood mononuclear cells, AIM = activation-induced markers, d = days, m = months.

S-specific CD4+ T-cells were detected in the majority of vaccinated individuals both early and late after vaccination, with relatively little waning over time (**Figure 3C, 3D**). Strikingly, CD4+ T-cell responses were consistently higher in mRNA-1273-vaccinated participants, although the BNT162b2-vaccinated cohort had too little power for comparison. We did not observe significant differences between CD4+ T-cell responses to WT, Delta, Beta or Omicron S peptide pools at either time point in any vaccination group (with the exception of a small but significant reduction to Delta and Omicron in the ChAdOx-1 S-vaccinated at 6 months, Friedman test with multiple comparisons). In general, a 1.2-to-1.9-fold reduction in T-cell responses to Omicron was observed (with the exception of the BNT162b2 group) (**Figure 3C, 3D**).

### Increased cross-reactivity of neutralizing antibodies and T-cells against Omicron after boost

Finally, we performed a preliminary analysis of neutralizing antibody and T-cell responses to the different SARS-CoV-2 variants after booster vaccination. HCW vaccinated with either one dose of Ad26.COV2.S (N=15) (*33*) or 2 doses mRNA-1273 (N=9) were included; both groups were boosted with BNT162b2, either at 85 days after primary vaccination (Ad26.COV2.S) or at 214 days after completion of the primary vaccination series (mRNA-1273) (**Figure 4A**). Specimen were collected 28 days (Ad26.COV2.S / BNT162b2) or 14 days (2x mRNA-1273/ BNT162b) after boost. Participant characteristics are described in **Table 2**.

**Figure 4.**
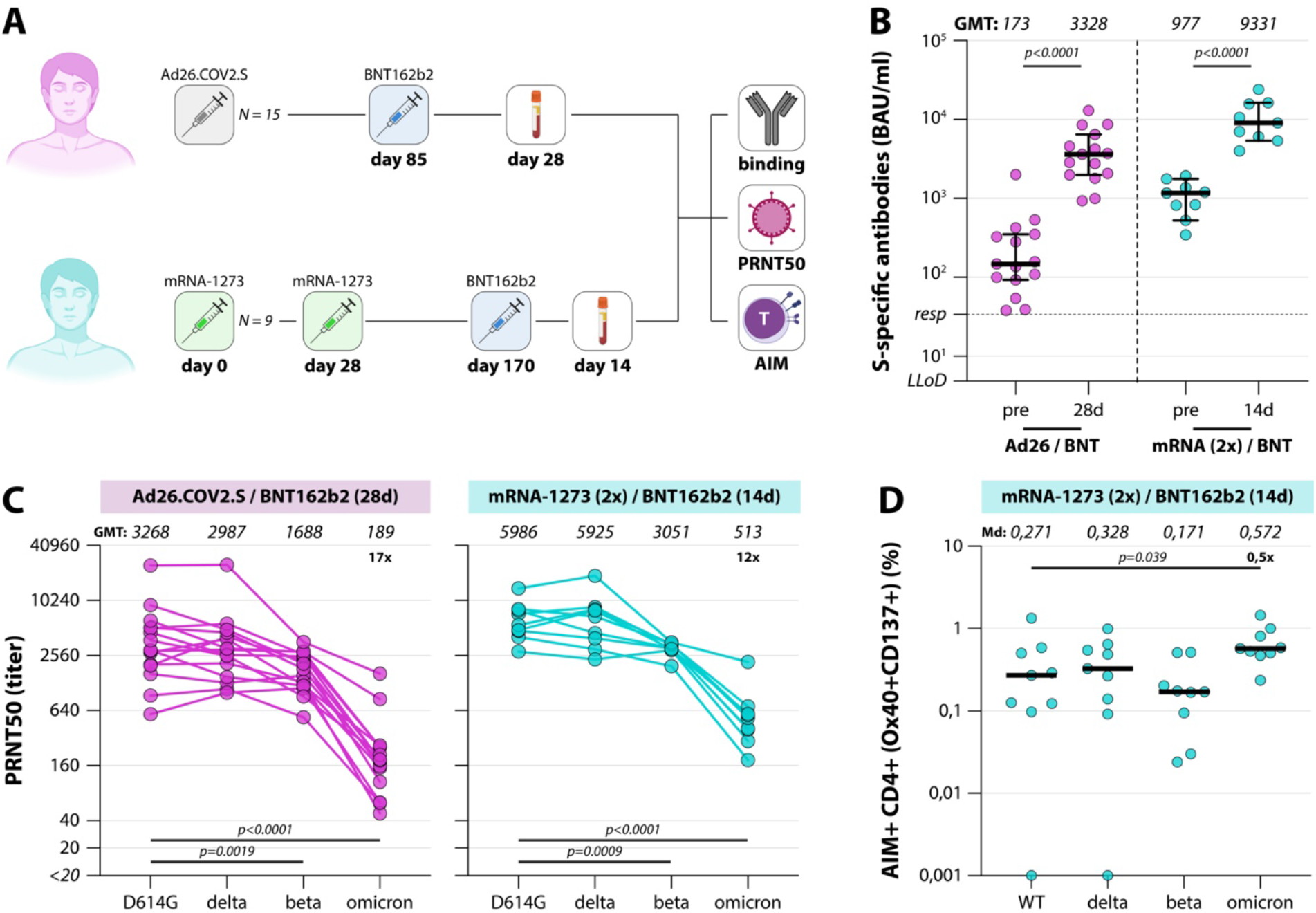
Variant-specific immune responses early booster vaccination. (A) Diagram of the number of included participants in the booster analysis. A total of N=24 participants were included for the analysis of binding antibodies, variant-specific neutralizing antibodies, and variant-specific T-cell responses. Responses were measured early after BNT162b2 booster vaccination. (B) Levels of binding S-specific antibodies early after booster vaccination. LLoD is 4.81 BAU/ml, responder (resp) cut-off is 33.8 BAU/ml (dotted line). Geometric mean titers are indicated above the graph. Comparisons of timepoints within study groups were performed by paired T test. (C) Levels of neutralizing antibodies in boosted donors. The lowest serum dilution tested was 1:20, undetectable PRNT50 values (<20) were set at a PRNT50 of 10. Geometric mean titers are indicated above the graphs, fold change reductions are indicated and calculated by dividing the D614G GMT by the Omicron GMT. (D) Upregulation of AIM in boosted donors. Percentages indicate the percentage of CD4+OX40+CD137+ cells after subtraction of observed background in a DMSO stimulation. Geometric means or medians are indicated above the graphs, fold change reductions are indicated and calculated by dividing the WT GMT or Md by the Omicron GMT or Md. Comparisons of VOC-specific responses within study groups were performed by Friedman test with multiple comparisons. BAU = binding arbitrary units, GMT = geometric mean titer, Md = median, PRNT50 = plaque reduction neutralization titer – 50%, AIM = activation-induced markers, d = days, m = months.

**Table 2.**
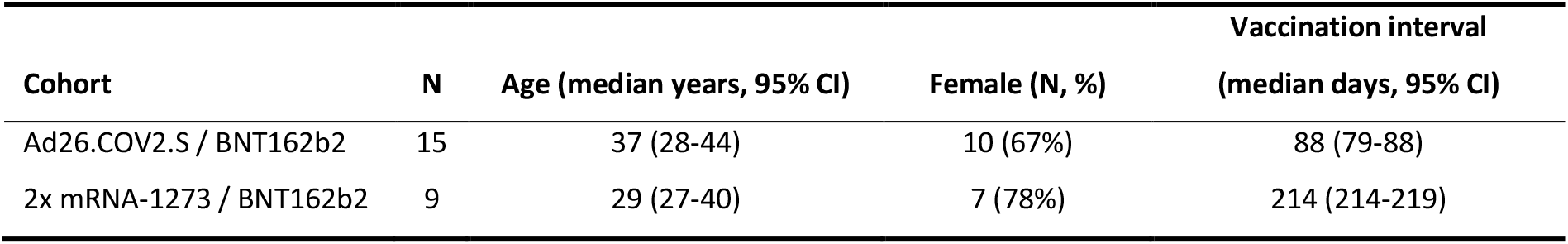
Booster participant characteristics.

| Cohort | N | Age (median years, 95% CI) | Female (N, %) | Vaccination interval<br>(median days, 95% CI) |
| --- | --- | --- | --- | --- |
| Ad26.COV2.S / BNT162b2 | 15 | 37 (28-44) | 10 (67%) | 88 (79-88) |
| 2x mRNA-1273 / BNT162b2 | 9 | 29 (27-40) | 7 (78%) | 214 (214-219) |

Both boosting strategies led to rapid recall responses and a significant increase in binding antibody levels was observed (**Figure 4B**); binding antibody levels in the mRNA-1273 boosting regimen reached the highest levels with a GMT of 9331 (versus 3328 in the Ad26.COV2.S-primed individuals). PRNT50 after boost were also higher in the mRNA-1273 primed individuals compared to the Ad26.COV2.S-primed individuals (**Figure 4C**). When comparing neutralization of the different VOC, no drop in neutralization of Delta, a two-fold drop in neutralization of Beta, but a significant 17- and 12-fold drop in neutralization of Omicron was still observed (p<0.0001 after Ad26.COV2.S priming, p=0.0002 after mRNA-1273 priming, Friedman test with multiple comparisons). However, neutralization of Omicron was observed or restored in all boosted individuals (GMT PRNT50 of 189 for Ad26.COV2.S-primed participants, 513 for mRNA-1273-primed individuals). An increase in S-specific T-cells was also observed after booster vaccination of mRNA-1273-primed participants. Interestingly, a 2-fold increase in T-cell responses to Omicron was observed when compared to WT, although this analysis was not powered with 9 participants (**Figure 4D**).

## Discussion

Here, we demonstrate the resistance of the SARS-CoV-2 Omicron (B.1.1.529) variant to neutralizing antibodies induced by mRNA-based or adenovirus vector-based vaccination. We observed up to 30-fold reductions of Omicron neutralizing titers when compared to WT SARS-CoV-2. Lower neutralizing antibody levels were previously associated with an increased risk of COVID-19 infection (*15*), and a higher burden of disease. In contrast, we show that SARS-CoV-2-specific CD4+ T-cell responses were minimally affected by mutations in the Omicron S protein. T-cell activity is thought to confer protection from severe disease, but it remains to be determined whether this is sufficient in the absence of a potent neutralizing antibody response. Importantly, a single BNT162b2 booster immunization induced a substantial increase in Omicron-specific neutralization and CD4+ T cells after priming with either Ad26.COV2.S or mRNA-1273. Neutralization of Omicron was partially restored in individuals without cross-reactive neutralizing antibodies prior to boosting.

We observed high, but transient, levels of SARS-CoV-2-specific binding antibodies after mRNA-based vaccination. Despite the waning of binding antibody levels in vaccinees, affinity maturation of antibodies and a continued increase in SARS-CoV-2-specific memory B cells was previously described between 3 and 6 months after mRNA vaccination (*34*). These memory B cells are generated in prolonged germinal center reactions (*35*), and are capable of mounting a rapid recall response upon re-encountering the S protein, either by natural infection or booster vaccination. In contrast, we observed that Ad26.COV2.S vaccination led to antibody responses which showed minimal waning at 6 months (*36*). This suggests that upon Ad26.COV2.S vaccination maturation of B cells occurs without further short-term boosting. Neutralizing antibodies are regarded the main correlate of protection against infection with SARS CoV-2 (*14, 15*). The capacity of sera from vaccinated individuals to neutralize the Omicron variant was reduced up to 30-fold, which is in line with another recent study (*37*). To be protected against symptomatic COVID-19 caused by Omicron, affinity maturation will be essential for the acquisition of broader neutralizing activity of RBD-binding antibodies that were previously formed (*38, 39*). Booster vaccination of Ad26.COV2.S- and mRNA-1273-primed individuals quickly increased S-specific antibodies and restored Omicron neutralization, however Omicron-specific levels were still significantly lower than neutralization of WT. Whether booster vaccinations increase the breadth of the response remains to be determined (*36*).

Multiple assays to determine the neutralizing capacity of sera are currently employed (*40*), and the variety of protocols hinders a direct comparison of different studies. We studied VOC-specific neutralization using infectious virus plaque reduction assays. It is known that SARS-CoV-2 propagation on VeroE6 cells, a cell-line routinely used for SARS-CoV-2 neutralization assays, can lead to mutations or deletions in the multibasic cleavage site in the S protein (*29, 31, 32*). We therefore grew the viral stocks and performed the neutralization assays on the TMPRSS2 expressing human airway cell-line Calu-3.

Thus far, little is known on cross-reactivity of T-cell responses with the Omicron variant(*20*). Previous studies into cross-reactivity of vaccine-induced T-cells with the Alpha, Beta, Gamma and Delta variant demonstrated minimal reductions in frequency and magnitude of T-cell responses to VOC (*8, 18, 19, 34, 41, 42*). Here, we performed a comprehensive analysis of SARS-CoV-2-specific CD4+ T-cells from vaccinated individuals early and late after receiving ChAdOx-1 S, Ad26.COV2.S, mRNA-1273, or BNT162b2 vaccination. In contrast to the humoral responses, we identified durable cellular immune responses that persisted for at least 6 months after either mRNA-based or adenovirus vector-based vaccination. Similar to observations on the level of binding antibodies, mRNA-1273 induced the highest level and most durable SARS-CoV-2-specific T-cell responses. Due to limited availability of PBMC from BNT162b2-vaccinated participants, only 5 donors instead of 15 in the other groups were included, so these results have to be interpreted with caution. However, in line with previous studies into VOC-specific T-cell responses, we show that vaccine-induced CD4+ T-cell responses equally recognize the WT, Beta, Delta, and Omicron variant independent of timing or vaccination regimen. Similar analyses have to be performed for CD8+ T-cells, however initial data indicated that these also retain reactivity (*20*). This fits well with the observation that although Omicron has numerous mutations in the S protein, T-cell epitopes are only minimally affected. A homologous or heterologous booster vaccination with BNT162b2 further increased T-cell responses.

Although this study is skewed towards healthy young-adult participants (with the exception of ChAdOx-1 S-vaccinated individuals) and in-depth analyses were performed in a relatively small number of subjects, we show that vaccinated individuals retain T-cell immunity to the Omicron variant, potentially balancing the lack of neutralization antibodies in preventing or limiting severe COVID-19. Furthermore, the fact that booster vaccinations restored and increased the neutralizing capacity against Omicron supports the call for immediate booster campaigns. In the near future, variant-specific booster vaccines may be required to optimally skew the immune responses towards emerging viruses.

## Data Availability

All data produced in the present study are available upon reasonable request to the authors.

## Acknowledgements

Chris Groen, Willemijn van der Kleij, Eveline de Haan, Roos Robert, virorunners and colleagues of the Erasmus MC HCW study team are acknowledged for their excellent study support. Faye de Wilt, Djenolan van Mourik, Suzanne van Efferen, Suzanne Hendrickx, Felicity Chandler, and Sandra Scherbeijn are thanked for their technical support and Chantal Reusken for providing the clinical specimen for SARS-CoV-2 Omicron culture. We acknowledge QIAGEN for supporting the study by providing the QuantiFERON SARS-CoV-2 RUO Starter Packs and Extended Packs. QIAGEN had no role in study design, data acquisition and analysis.

## Supplementary Material

### Methods

#### Ethics statement

Samples from three different trials (HCW, ConCOVID, and SWITCH) were analysed in the scope of this study (**Supplemental Table 1**). The HCW study was approved by the institutional review board of the Erasmus MC (medical ethical committee, MEC-2020-0264) (*1*). The ConCOVID trial was also approved by the institutional review board of the Erasmus MC (MEC 2020-0228). The study was registered at clinicaltrials.gov (NCT04342182) (*2*). The SWITCH trial was approved by the institutional review board of the Erasmus MC (MEC 2021-0132) and local review boards of participating centres. The study was registered at clinicaltrials.gov (NCT04927936) (*3*). All studies adhere to the principles of the Declaration of Helsinki, and written informed consent was obtained from every participant, patient or legal representative.

#### Study design

A prospective cohort study of health care workers (HCW) at Erasmus MC was initiated in 2020, with a focus on immunity against SARS CoV-2 after symptomatic presentation to the occupational health services. From January 2021, HCW in the Netherlands were offered vaccination with one of the 4 approved vaccines in the Netherlands (ChAdOx-1 S, Ad26.COV2.S, mRNA-1273 and BNT162b2), participants of the HCW study were invited for the follow up study in which immune responses upon vaccination were studied. From December 2021, HCW in the study were invited for a follow up study after booster vaccination with BNT162b2. Serum and PBMC samples from all vaccinated participants of the HCW study were obtained 28 days and 6 months after second vaccination (with the exception of Ad26.COV2.S vaccinated HCW, samples were obtained 56 days after first vaccination). HCW were vaccinated with (1) ChAdOx-1 S / ChAdOx-1S (56 day interval), (2) Ad26.COV2.S, (3) mRNA-1273 / mRNA-1273 (28 day interval), or BNT162b2 / BNT162b2 (21 days interval) (**Figure 1A**). Additional serum and PBMC samples were obtained from 9 mRNA1273 / mRNA-1273-primed participants at 14 days after booster vaccination with BNT162b2. Participants of the SWITCH trial were vaccinated with Ad26.COV2.S, followed by a BNT162b2 boost (85 day interval) (*3*). Serum samples were obtained 28 days after the booster vaccination (**Figure 4A**). Participants of the HCW and SWITCH trial had no history of previous SARS-CoV-2 infection, as confirmed by absence of N-specific antibodies and / or S-specific antibodies pre-vaccination at baseline. Serum samples from convalescent PCR-confirmed COVID-19 patients were obtained 6 months after infection in the scope of the ConCOVID study, in which study participants were recruited to participate as plasma donors (*2*). These patients were infected during the first wave in 2020, all with a D614G SARS-CoV-2. In-depth neutralization and T-cell assays were performed on a selection of N=15 participants from each group (**Table 1, Table 2**). The selection of participants for in-depth analyses was based on availability of longitudinal PBMC samples and the distribution of antibody responses (**Figure 1B, 1C**).

#### Serum and PBMC isolation

Serum was collected in 10-ml tubes without anticoagulant, centrifuged at 2500 rpm for 15 min, aliquoted, and stored at −20°C for further experiments. PBMCs were isolated from blood collected in K_3_EDTA or Lithium Heparin tubes by density gradient centrifugation. Briefly, blood was layered on a density gradient (Lymphoprep, STEMCELL Technologies), and PBMCs were separated by centrifuging at 2000 rpm for 30 min. PBMCs were washed four times in PBS and subsequently frozen in liquid nitrogen in 90% fetal bovine serum (FBS) with 10% dimethyl sulfoxide (DMSO; Honeywell) until use in stimulation assays.

#### Detection of S-specific binding antibodies

Binding antibodies against the SARS-CoV-2 Spike (S) protein were measured by Liaison SARS-CoV-2 TrimericS IgG assay (DiaSorin, Italy), with a lower limit of detection of 4.81 BAU/ml and a cut-off for positivity at 33.8 BAU/ml. The assay was performed following the manufacturer’s instructions.

#### IFNγ release assay (IGRA)

The SARS-CoV-2-specific T cell response was measured by commercially available IFNγ Release Assay (IGRA, QuantiFERON, Qiagen) in whole blood as previously described and following the manufacturer’s description (*4*). In short, heparinized whole blood was incubated with three different SARS-CoV-2 antigens for 20-24h using a combination of peptides stimulating both CD4^+^ and CD8^+^ T-cells (Ag1, Ag2, Ag3, QuantiFERON, QIAGEN). After incubation, plasma was obtained and IFNγ production in response to the antigens was measured by ELISA. Results are expressed in IU IFNγ /ml after subtraction of the NIL control values as interpolated from a standard calibration curve. Lower limit of detection in this assay is set at 0.01 IU/ml, responder cut-off is 0.15 IU/m). IFNγ production after stimulation with Ag2, containing peptides covering the S protein, was shown in this study.

#### Virus culture and deep-sequencing

SARS-CoV-2 isolates were grown to passage 3 on Calu-3 (ATCC HTB-55) cells in Advanced DMEM/F12 (Gibco), supplemented with HEPES, Glutamax, penicillin (100 IU/mL) and streptomycin (100 IU/mL) at 37°C in a humidified CO_2_ incubator. Infections were performed at a multiplicity of infection (MOI) of 0.01 and virus was harvested after 72 hours. The culture supernatant was cleared by centrifugation at 1000 x g for 5 min and stored at −80°C in aliquots. Stock titers were determined by incubating 10-fold dilutions of virus stock in OptiMEM at 37°C for 1 hour, after which they were transferred onto Calu-3 cells and incubated for 8 hours at 37°C in a humidified CO_2_ incubator. Next, cells were fixed with 4% formalin, permeabilized in 70% ethanol, after which infected cells were stained with polyclonal rabbit anti–SARS-CoV-2 nucleocapsid antibody (Sino Biological) and a secondary goat anti-rabbit IgG AF488 (Invitrogen). Stained plates were scanned on the Amersham Typhoon Biomolecular Imager (channel Cy2; resolution 10 µm; GE Healthcare) and the number of infected cells was determined to calculate the stock titer per milliliter. All work with infectious SARS-CoV-2 was performed in a Class II Biosafety Cabinet under BSL-3 conditions at Erasmus Medical Center.

Viral genome sequences were determined using Illumina deep-sequencing. RNA was extracted using AMPure XP beads and cDNA was generated using ProtoscriptII reverse transcriptase enzyme (New England, BiotechnologyBioLabs) according to the manufacturer’s protocol (*5*). Samples were amplified using the QIAseq SARS-CoV-2 Primer Panel (Qiagen). Amplicons were purified with 0.8x AMPure XP beads and 100ng of DNA was converted to paired-end Illumina sequencing libraries using the KAPA HyperPlus library preparation kit (Roche) with the KAPA unique dual-indexed adapters (Roche) as per manufacturers recommendations. The barcode-labeled samples were pooled and analyzed on an Illumina sequencer V3 MiSeq flowcell (2×300 cycles). Sequences were analyzed using CLC Genomics Workbench 21.0.3. The 614G virus (clade B; isolate Bavpat-1; European Virus Archive Global #026 V-03883) passage 3 sequence was identical to the passage 1 (kindly provided by Dr. Christian Drosten) and no minor variants >20% were detected. The beta variant (clade B.1.351) passage 3 sequence contained two mutations compared the original respiratory specimen: one synonymous mutations C13860T (Wuhan-1 position) in ORF1ab and a L71P change in the E gene (T26456C, Wuhan-1 position). No other minor variants >20% were detected. The delta (clade B.1.617.2) and omicron (clade B.1.1.529) variant passage 3 sequences were identical to the original respiratory specimens and no minor variants >20% were detected. Due to primer mismatches in the S1 region of the omicron spike gene, amplicons 72, 73, 75 and 76 were sequenced at low coverage. Therefore, the S1 regions of the original respiratory specimen and passage 3 virus were confirmed to be identical by Sanger sequencing. The beta, delta and omicron sequences have been submitted to Genbank.

The beta variant contained the following spike changes: L18F, D80A, D215G, del241-243, K417N, E484K, N501Y, D614G, and A701V. The delta variant contained the following spike changes: T19R, G142D, del156-157, R158G, A222V, L452R, T478K, D614G, P681R and D950N. The omicron variant contained the following spike mutations: A67VS, del69-70, T95I, G142-, del143-144, Y145D, del211, L212I, ins215EPE, G339D, S371L, S373P, S375F, K417N, N440K, G446S, S477N, T478K, E484A, Q493R, G496S, Q498R, N501Y, Y505H, T547K, D614G, H655Y, N679K, P681H, N764K, D796Y, N856K, Q954H, N969K, L981F.

#### Detection of neutralizing antibodies by plaque reduction assay

Plaque reduction neutralization tests (PRNT) were performed as described previously (*1, 6*). Briefly, heat-inactivated sera were two-fold diluted in OptiMEM medium starting at a dilution of 1:10 (or 1:80 for sera known to have more than 2500 BAU/ml S-specific binding antibodies) in 60ul. 400 plaque forming units of different SARS-CoV-2 variants were added to each well in 60ul of virus suspension incubated at 37°C for 1 hour. After 1 hour of incubation, the virus-antibody mixtures were transferred onto the human airway cell line Calu-3 and incubated for 8 hours. After incubation, cells were fixed and plaques were stained with polyclonal rabbit anti–SARS-CoV-2 nucleocapsid antibody (Sino Biological) and a secondary peroxidase-labeled goat anti-rabbit IgG (Dako). Signal was developed by using a precipitate-forming 3,3’,5,5’-tetramethylbenzidine substrate (TrueBlue; Kirkegaard & Perry Laboratories) and the number of infected cells was counted per well by using an ImmunoSpot Image Analyzer (CTL Europe GmbH). The dilution that would yield 50% reduction of plaques (PRNT50) compared with the infection control was estimated by determining the proportionate distance between two dilutions from which an endpoint titer was calculated. Raw data for neutralizing antibodies early after vaccination (**Supplemental Figure 2A, B**), late after vaccination (**Supplemental Figure 2C, D**), 6 months after positive PCR (**Supplemental Figure 2E, 2F**), or early after booster vaccination (**Supplemental Figure 2G, H**) are included in the supplemental figures. Infection controls and positive serum controls were included on each plate, the NIBSC standard was included in three separate experiments. When no neutralization was observed, we set the PRNT50 one dilution step below the dilution series, i.e., a PRNT50 value of 10.

#### Overlapping SARS-CoV-2 Spike peptide pools

SARS-CoV-2 peptides were synthesized as crude material (TC Peptide Lab, San Diego, CA). Overlapping 15-mer by 10 amino acids covering the full-length S proteins from the WT, Beta (B.1.351), Delta (B.1.617.2) and Omicron (B.1.1.529) variants were synthesized and individually resuspended in dimethyl sulfoxide (DMSO) at a concentration of 10–20 mg/mL. Megapools (MP) for each spike variant were generated by pooling aliquots of these individual peptides, undergoing another lyophilization, and resuspending in DMSO at 1 mg/mL (*7*).

#### Stimulations for detection of SARS-CoV-2-specific T-cells

PBMCs were thawed in Gibco Roswell Park Memorial Institute 1640 medium (Gibco) supplemented with 10% human serum (Sanquin, Rotterdam), penicillin (100 IU/ml; Lonza, Belgium), streptomycin (100 μg/ml; Lonza, Belgium), and 2 mM L-glutamine (Lonza, Belgium; R10H medium) and treated with Benzonase (50 IU/ml; Merck) at 37°C for 30 min. Subsequently, 1 × 10^6^ PBMCs were stimulated with SARS-CoV-2 variant peptide pools at 1 μg/ml per peptide in 200 μl in a 96-well U-bottom plate at 37°C for 20 hours. Cells were additionally stimulated with an equimolar concentration of DMSO (negative control) or a combination of phorbol 12-myristate 13-acetate (50 μg/ml) and ionomycin (500 μg/ml) (positive control). Additionally, PBMC were stimulated with a peptide pool consisting of 176 known peptides for a broad range of HLA subtypes and different infectious agents (including human herpesviruses and influenza virus) as peptide positive control (PepMix CEFX Ultra SuperStim Pool, JPT). After stimulation, cells were stained for phenotypic lymphocyte markers.

#### Detection of AIM by flow cytometry

Detection of AIM was performed as described previously (*1*). Briefly, cells were stained with the following antibodies in their respective dilutions: anti–CD3-PerCP (1:25, clone SK7, BD), anti–CD4-V50 (1:50, clone L200; BD), anti–CD8–fluorescein isothiocyanate (1:25, clone DK25; Dako), anti–CD45RA-phycoerythrin (PE)–Cy7 (1:50, clone L48; BD), anti–CCR7-BV711, anti–CD69-allophycocyanin (APC)-H7 (1:50, clone FN50; BD), anti–CD137-PE (1:50, clone 4B4-1; Miltenyi), and anti–OX40-BV605 (1:25, clone L106; BD). LIVE/DEAD Fixable Aqua Dead Cell staining was included (1:100, AmCyan; Invitrogen) and acquired on a FACSLyric (BD Biosciences). SARS-CoV-2–specific T-cells were identified by gating the CD4^+^Ox40^+^CD137^+^ and CD8^+^CD69^+^CD137^+^ cells. The DMSO-stimulated sample was used to set the cutoff gate for activation markers. On average, 500,000 cells were acquired per sample.

#### Statistical analysis

A comparison of the baseline characteristics age and interval between vaccination between groups was performed by Kruskal-Wallis with multiple comparisons. Reduction in binding and neutralizing antibody levels over time within groups was analyzed by paired T test. Different groups were compared by unpaired T test. Differences in PRNT50 and T-cell responses to VOC were estimated by Friedman test with multiple comparisons, data was not normally distributed. Spearman R was calculated for the correlation between binding and neutralizing antibodies. All statistical analyses were performed on log-transformed data.

#### Software

Statistical analyses were performed with Graphpad PRISM version 9.1.2. Expression of AIM was analysed with FlowJo software version 10.8.1.

## Supplemental Tables

**Supplemental Table 1.**
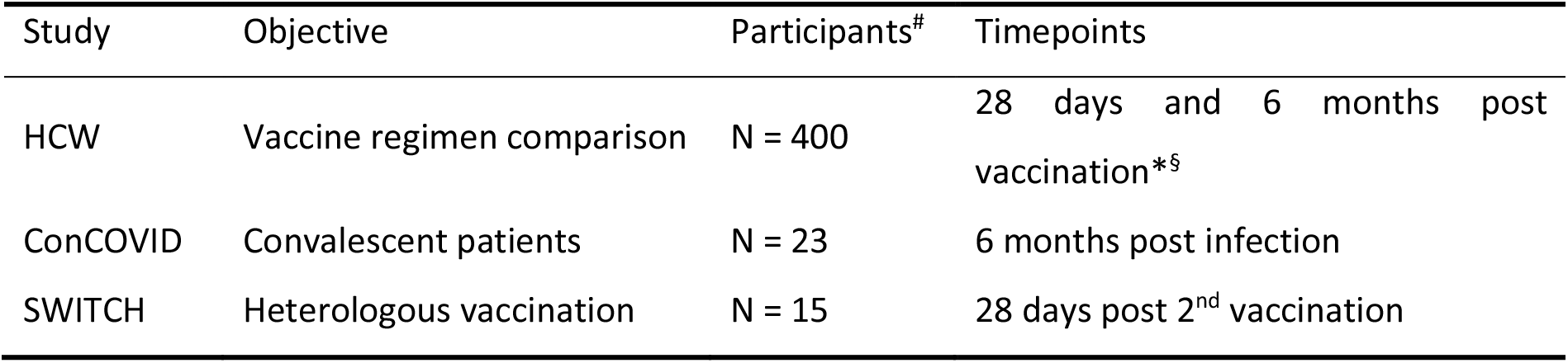
Cohort background. A total of 438 participants from three different trials (HCW, ConCOVID, SWITCH) were included in this study. * N = 9 participants of the HCW study were additionally analysed 14 days after the 3^rd^ vaccination. ^§^ Ad26.COV2.S samples were analysed 56 days and 6 months after 1^st^ vaccination. ^#^ T-cell responses were exclusively assessed in participants of the HCW study for vaccine regimen comparisons.

| Study | Objective | Participants<sup>#</sup> | Timepoints |
| --- | --- | --- | --- |
| HCW | Vaccine regimen comparison | N = 400 | 28 days and 6 months post vaccination*§ |
| ConCOVID | Convalescent patients | N = 23 | 6 months post infection |
| SWITCH | Heterologous vaccination | N = 15 | 28 days post 2<sup>nd</sup> vaccination |

## Supplemental Figures

**Supplemental Figure 1.**
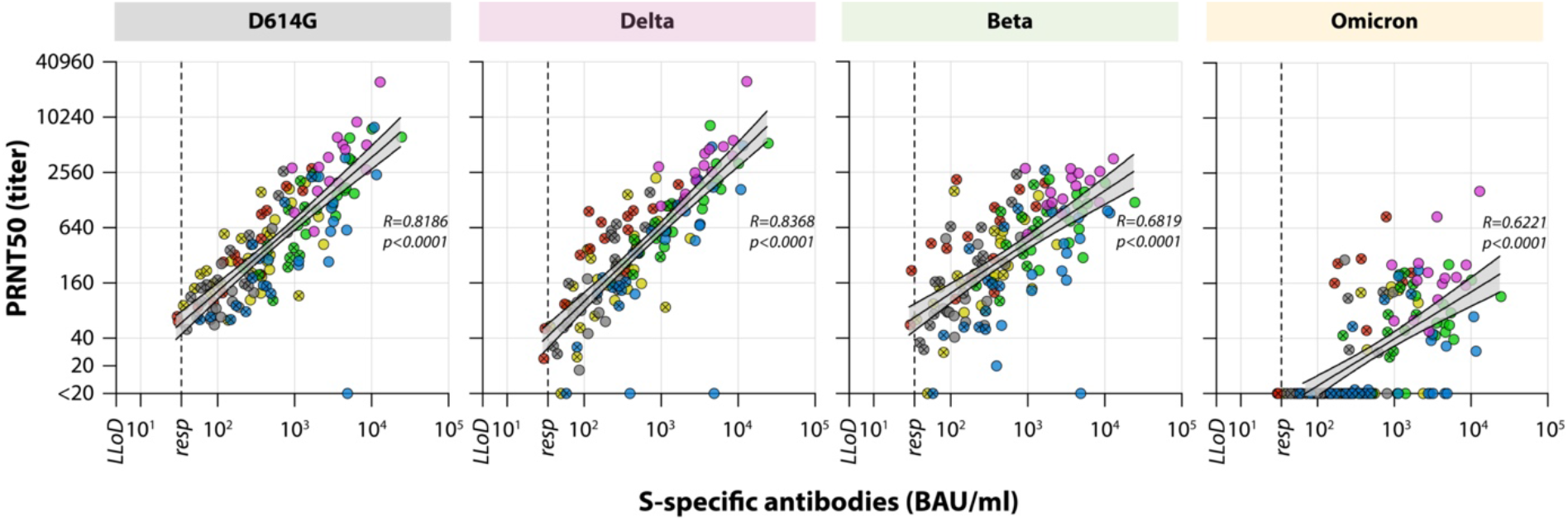
Correlation between neutralizing antibodies and S-specific binding antibodies. Correlation between S-specific biding antibodies early (open symbols) and late (symbols with cross) after vaccination or infection with D614G-specific, Delta-specific, Beta-specific, or Omicron-specific neutralizing antibodies. For S-specific binding antibodies the LLoD is 4.81 BAU/ml, responder (resp) cut-off is 33.8 BAU/ml (dotted line). For neutralizing antibodies, the lowest serum dilution tested was 1:20, undetectable PRNT50 values (<20) were set at a PRNT50 of 10. LLoD = lower limit of detection, BAU = binding arbitrary units, PRNT50 = plaque reduction neutralization titer – 50%. Spearman R was calculated on basis of log-transformed data.

**Supplemental Figure 2.**
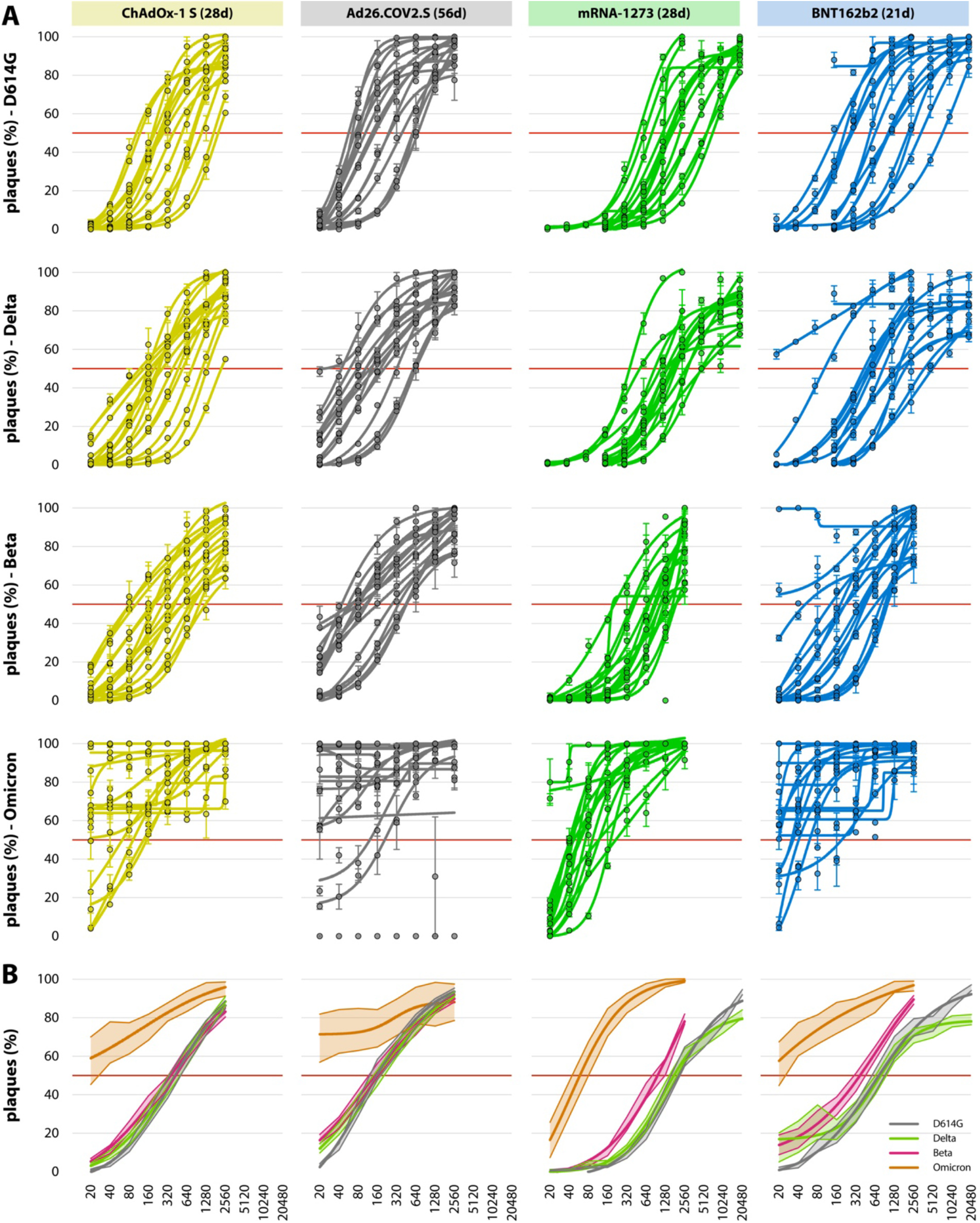
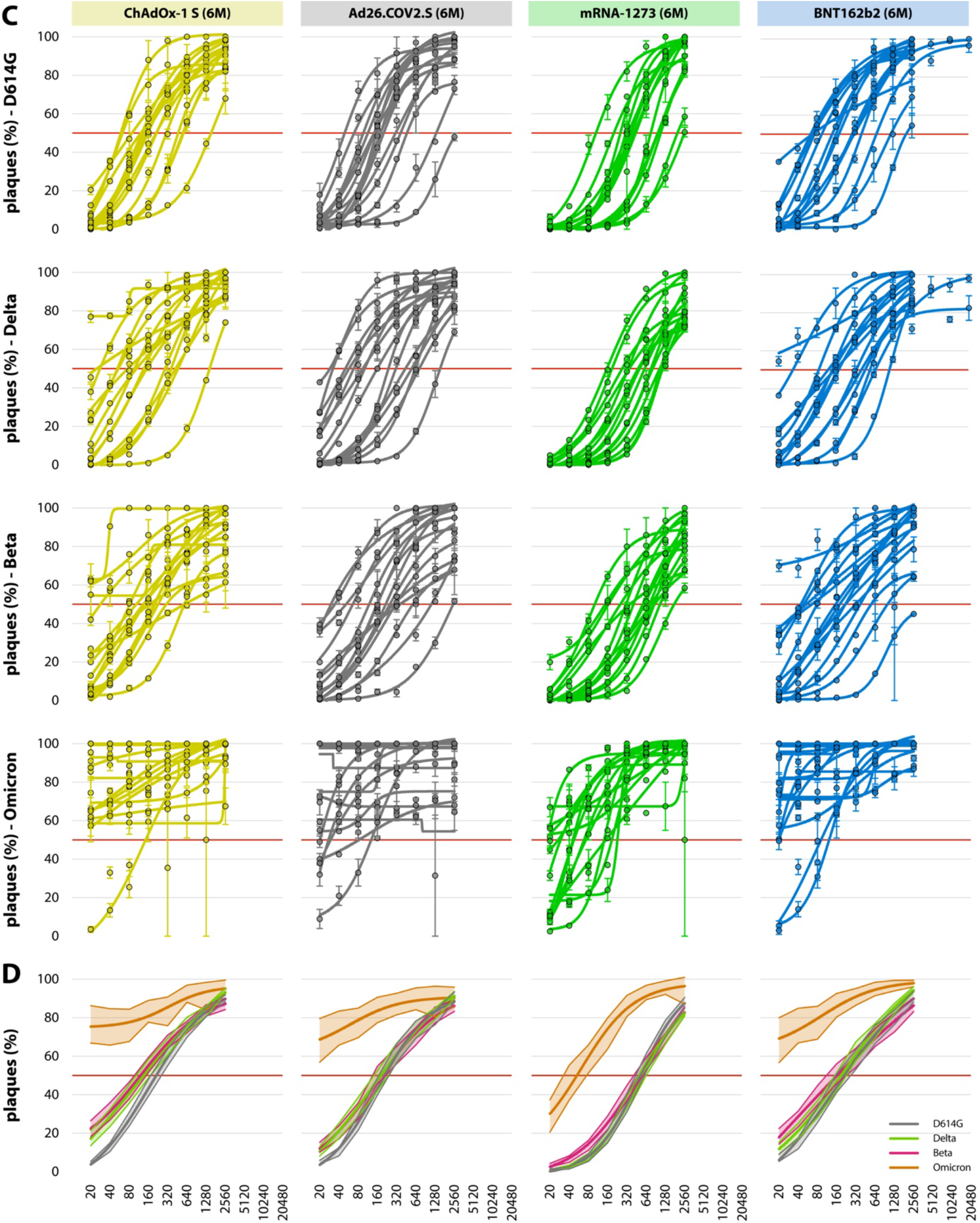
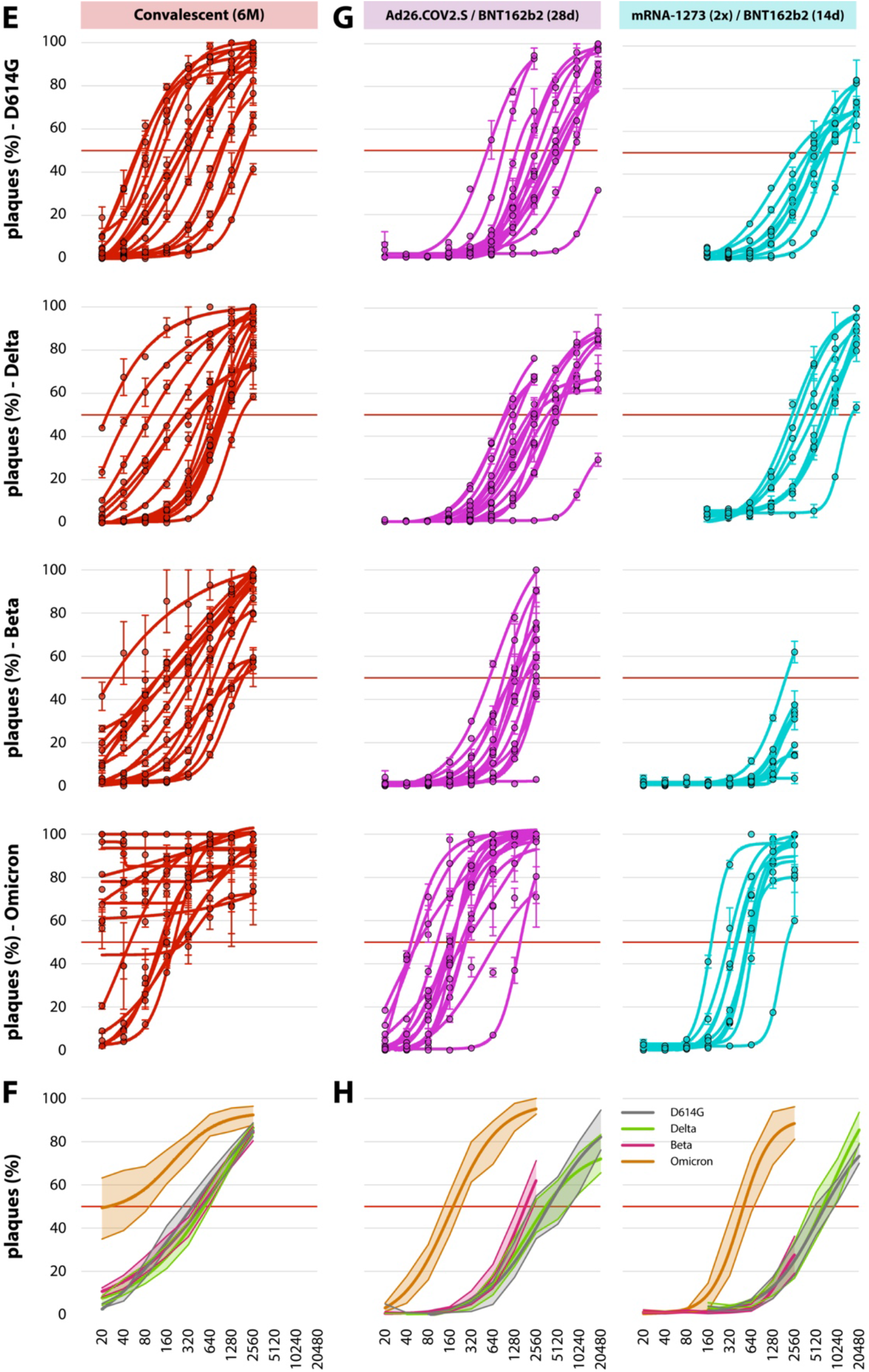
PRNT50 calculations. Graphs in panels A, C, E, and G show per participant log(inhibitor) versus responses curves with four parameter variable slopes based on plaque counts compared to the virus control, for the four different variants. Panels B, D, F, and H show the average curves from all participants with the 95% CI. Neutralizing antibodies (A,B) early after vaccination, (C,D) late after vaccination, (E,F) 6 months after infection, and (G,H) early after booster vaccination.

